# Longitudinal associations between parental bonding and child preschool social-emotional problems: The unique and combined role of mothers and fathers

**DOI:** 10.1101/2025.01.30.25321411

**Authors:** Enni Hatakka, Marjo Flykt, Erja Rusanen, Anneli Kylliäinen, E. Juulia Paavonen, Olli Kiviruusu

## Abstract

Research is currently limited on the impact of maternal and especially paternal postpartum bonding on child social-emotional development. This study is part of the nationally representative CHILD-SLEEP -sample (n = 710 families), where postpartum bonding was assessed with Postpartum Bonding Questionnaire (PBQ) at the child age of eight months, and child social-emotional problems were assessed with the Five to Fifteen (FTF) and the Strengths and Difficulties Questionnaire (SDQ) at child age of five years. Both maternal and paternal postpartum bonding difficulties at eight months were associated with child externalizing, internalizing and peer problems at five years. When parental depressive symptoms were controlled, the association remained significant only with internalizing problems. When investigating joint maternal and paternal bonding, we found both cumulative and mother-driven effects with child internalizing problems. The results emphasize the role of early parental bonding for later child mental health, highlighting the importance of early parental support.

## Introduction

The quality of early parent-child interaction plays a significant role in children’s social-emotional development. Postpartum bonding refers to the emotional bond parents develop towards their baby, beginning already during pregnancy and further evolving through interactions with the baby (Condon & Corkindale, 1998). Recent studies have shown an association between maternal bonding difficulties and infant and toddler social-emotional symptoms (LeBas et al., 2019; LeBas et al., 2022; Murakami et al., 2023; Rusanen et al., 2022), but more research is needed including both mothers and fathers with longer follow-ups.

Condon & Corkindale (1998) have defined postpartum bonding as comprising caregiver’s feelings of competence in meeting their child’s needs, subjective pleasure derived from the interaction with the child as well as acceptance of the parental role and its demands. According to Condon (1993), bonding consists of two dimensions: *intensity* and *quality.* Intensity refers to the amount of time and energy parents spend mentally occupied with their child, while quality entails the perceived pleasure, closeness and tenderness in the parent-child interaction. Both mothers and fathers have the ability to develop mental representations and an emotional bond towards their yet unborn child, right from the start of the pregnancy (Vreeswijk et al., 2014; Vreeswijk et al., 2015). Difficulties in the prenatal bond have been found to be associated with postpartum bonding difficulties (Rusanen et al., 2021). After birth, the interactions with the newborn child further develop and modify parental bonding. On behavioral level, postpartum bonding manifests through interactional aspects such as sensitivity to the child’s needs (Le Bas et al., 2022). Problematic postpartum bonding can manifest in early interaction as parental anger, anxiety, and rejection towards the child (Brockington et al, 2006). Conversely, the child reads the parent’s emotional signals, which in turn affect the child’s attachment behavior. Attachment-related behaviors include the emotional tone of interacting with the parent and seeking their physical proximity when under distress, (e.g., Cadman et al., 2018). This way, difficulties in bonding may lead to a negative loop of dysfunctional interactions between the parent and the child and affect more broadly on child’s social-emotional development. Attachment behavior and its association with later child development and psychopathology has been widely studied, but there is significantly less studies on the effect of parental bonding on later child development.

Social-emotional development is a central area of development, with roots in early childhood. Developmental problems in this area include internalizing and externalizing problems as well as difficulties in interacting with peers. Externalizing behavior depicts outwards-directed maladaptive behavior, such as aggression (Campbell et al., 2000; Liu, 2004). Internalizing behavior refers to withdrawal behavior, with the focus on the child’s internal experience (Liu, 2004) around the feelings of anxiety and depression (Stacks, 2005). Problems in peer relations in early childhood include bullying and exclusion. Bullied preschool-aged children suffer from feelings of loneliness and are in a risk of long-term negative consequences such as adolescent internalizing problems and substance use (Hay et al., 2004; Mesman et al., 2001; see for a meta-analysis, Moore et al., 2017).

Previous research indicates that maternal bonding affects child development in several ways, although most research has concerned prenatal bonding (de Waal et al., 2023; see for a meta-analysis, Le Bas et al., 2020; Le Bas et al., 2022; Murakami et al., 2023; Rusanen et al., 2022). In a meta-analysis including mostly measures of prenatal bonding, higher maternal bonding quality was found to have a positive impact on infant attachment security and mood, and a negative impact on temperament difficulty (Le Bas et al., 2020, 2022). Also, recent studies including both pre- and postpartum bonding have found an association between difficulties in maternal bonding and children’s social-emotional problems at the age of two to four (de Waal et al., 2023; Le Bas et al., 2022; Rusanen et al., 2022). To better understand the long-term effects of parental bonding on child social-emotional development, this study extends the follow-up period until the child age of five years.

Earlier postpartum bonding research has exclusively focused on mothers, which does not correspond with the family systems of the present day, with increased paternal caregiving role. Research has highlighted that the father-child-relationship can also have a strong impact on child wellbeing and development (e.g., Fonseca et al., 2018; Verschueren & Marcoen, 1999), but the role of paternal bonding is still unclear. A recent study on maternal and paternal postpartum bonding showed that paternal bonding predicted the quality of maternal bonding (Bieleninik et al., 2021). While there are yet no studies combining maternal and paternal bonding, studies on other related concepts such as child attachment indicate that children who are insecurely attached to two parents suffer more from social-emotional problems than children securely attached to one parent (e.g. Kochanska & Kim, 2013). These findings support the importance of studying joint parental bonding quality in two-parent families, which we aim to investigate in the current study.

In the recent postpartum bonding studies, maternal bonding quality itself was associated with child social-emotional problems regardless of the level of maternal depressive symptoms (Le Bas et al., 2022; Murakami et al., 2023; Rusanen et al., 2022). Yet, depressive symptoms have generally been linked with more problematic pre- and postpartum bonding (Brockington et al., 2006; McNamara et al., 2019; Rusanen et al., 2018), as depression may weaken parental emotional involvement with their child (Figueiredo & Costa, 2009). Thus, it is important to consider the role of parental depressive symptoms when investigating parental postpartum bonding.

In this study, we investigated whether maternal bonding difficulties at eight months are associated with child’s social-emotional problems (internalizing, externalizing, and peer problems) at five years. Based on previous bonding studies with shorter follow-up time (e.g., Murakami et al., 2023; Rusanen et al., 2022), we hypothesized that maternal bonding difficulties would be associated with child’s social-emotional problems. Secondly, we aimed to examine whether paternal bonding difficulties at eight months are associated with the same child’s social-emotional problems at five years of age. Even without prior studies on paternal bonding, we assume that also fathers’ difficulties in bonding would be associated with child’s social-emotional problems, as for example paternal mental well-being has been shown to affect child development (e.g., Ramchandani et al., 2008). We also examined the joint effect of both parents’ bonding quality on child social-emotional problems. Based on attachment studies (e.g., Kochanska & Kim, 2013), we hypothesized a cumulative effect, i.e., that bonding difficulties in both parents would lead to more social-emotional problems than problems in only one parent. Lastly, we investigated how controlling for parental depressive symptoms affects the association between parental postpartum bonding and child social-emotional problems. Our hypothesis was that controlling for parental depressive symptoms would not change the association between parental postpartum bonding and child social-emotional problems.

## Methods

### Participants and procedure

The current study is a part of a longitudinal CHILD-SLEEP-cohort study by the Finnish Institute for Health and Welfare consortium including Tampere university hospital, Tampere University, University of Eastern Finland and University of Helsinki. The main aim of the cohort study is to investigate children’s quality of sleep, development, and health (Paavonen et al., 2017). The study protocol was approved by Pirkanmaa Hospital District Ethical Committee (9.3.2011, ethical research permission code R11032). The general population sample consisted of parents from 63 maternity clinics of Pirkanmaa Hospital District in central Finland between April 2011 and December 2012. The parents were informed about the study during their regular visit to the maternity clinic, approximately at the 32^nd^ gestational week. Only Finnish-speaking families were considered eligible as the questionnaires were available in Finnish only. The parents willing to participate in the study were given consent forms and then the prenatal questionnaires to fill (N = 2244 families), and a total of 1673 families returned the baseline questionnaires and the consent forms. In addition to the third trimester of pregnancy (baseline), measurements took place at 3, 8, 18, 24 months, and 5 years postpartum.

The sample for this study included families who took part in the prenatal, 8-month and 5-year time points. To be included in the present study, one of the parents had to answer at least the postpartum bonding questionnaire at eighth months and one of the two 5-year follow-up questionnaires. In families with twins (n=4), answers concerning only one twin were included in this study. The 5-year questionnaires were either answered by mothers alone (two thirds of the families) or both parents together (one third). There was a 41% total follow-up rate from the prenatal sample to the 5-year follow up sample used in this study. A total of 710 families were included in the current study, with answers from 708 mothers and 681 fathers, concerning their children (337 girls and 373 boys).

Attrition analyses, comparing the included 710 families to all baseline participants were conducted on the following background factors: education level, parental age, number of children in the family, and parental depressive symptoms. The only significant associations were that in the families taking part in the 8-month and 5-year follow up, mothers’ mean age was higher compared to families who did not continue in the 5-year wave. Also, parents taking part in the 8-month and 5-year follow-up had less children than parents at baseline.

### Measures

#### Postpartum bonding

Parental postpartum bonding at eight months was measured with the Postpartum Bonding Questionnaire (PBQ), a widely used scale with good psychometric properties (Brockington et al., 2006; Van Bussel et al., 2010). One 12-item subscale from the questionnaire was used, depicting a general “bonding problem factor” (Brockington et al, 2006). All the 12 items have six response options from 0 to 5, and a summary variable (range 0 – 60) is formed based on all items, representing problematic postpartum bonding (Brockington et al., 2006). The scale comprises items such as: “My baby irritates me”, and “I feel close to my baby” (reversed). Higher values on the scale thus indicate greater difficulties for the parent to form an emotional bond with the child. Internal consistency for maternal (α=.82) and paternal postpartum bonding (α=.79) was good. In the analyses the continuous scale score was used. To help interpretation of the interactions visually, the scale score was dichotomized using the general cutoff of ≥12 points for clinical bonding disorder (Brockington, 2006).

#### Child social-emotional problems

Child social-emotional problems at 5 years were measured using the Five to Fifteen (FTF, Kadesjö et al., 2004) and the Strengths and Difficulties Questionnaires (SDQ, Goodman, 1997). FTF is a Nordic questionnaire which was developed for screening psycho-social and cognitive development in children aged 5-15 (Kadesjö et al., 2004). The questionnaire has been translated to Finnish and has been found to have good to excellent internal consistency and good retest stability (Kadesjö et al., 2004). The scale includes 181 items that cover eight developmental domains, of which the current study used internalizing and externalizing problems. The questionnaire has a 3-point Likert scale ranging from “does not apply” (0), “applies sometimes or to some extent” (1), to “definitely applies” (2). The internalizing scale consists of 12 items concerning internalizing symptoms with a maximum score of 24, and the externalizing scale has 13 items with a maximum score of 26. The internalizing scale comprises items such as “appears tense and worried” and the externalizing scale consists of items such as “refuses to follow adult’s instructions”. In the current data, internal consistency varied from acceptable to very good (internalizing α = .59; externalizing α = .83).

SDQ is a widely used short measure of child mental health that covers five different developmental domains (Goodman, 1997), of which the current study used emotional and conduct problems, as well as problems in social relations with peers. The emotional problems scale includes items such as “often unhappy, downhearted or tearful”, the conduct problems scale has items such as “often has temper tantrums or hot tempers”, and the peer problems scale includes for example “picked on or bullied by other children”. All the three domains consist of five items with a 3-point Likert scale ranging from “not true” (0), “ somewhat true” (1), to “certainly true” (2). Previous studies have found satisfactory internal consistency for the questionnaire (e.g., Goodman. 2001), and in this data, internal consistency was somewhat acceptable (emotional problems α = .55; conduct problems α = .58; peer problems α = .47).

#### Background factors

In addition to child sex (available from the study database), several parental background factors were used as covariates in the statistical analyses, chosen based on theoretical assumptions of factors having an impact on early bonding and child development. Both the mother’s and the father’s age, the number of other children, and education level were reported in the first data collection wave during the third trimester of pregnancy. Parental education level was inquired using six categories: “no degrees”, “an occupational course/courses”, “occupational school”, “institute of applied sciences degree”, “university degree”, and “other”. For the analyses, a three-category variable was dummy coded: 1 – no degree/other; 2 – occupational course/courses/school or degree of applied sciences; 3 – university degree.

#### Depressive symptoms

Parental postpartum depressive symptoms at eight months were measured with the Center for Epidemiologic Studies Depression (CES-D) 10-item version (Radlof, 1977), where the suicidality question was left out for ethical reasons. The CES-D has been found to have good psychometric properties across different populations (Björgvinsson et al., 2013; Mohebbi et al., 2018). The 10-item version consists of response options varying from 0 (rarely/not at all/less than once a week) to 3 (all the time/5-7 times per week), with a maximum sum score of 24. In this data, there was good internal consistency for maternal postpartum depression (α = .80) and paternal postpartum bonding (α = .83).

### Statistical analysis

All analyses were conducted with SPSS 28.0 for Windows software. As a preliminary inspection, we ran descriptive statistics on the variables of interest. Outliers were found on two cases in maternal and paternal bonding variables and in one case on FTF internalizing scale. Winsorizing technique was used to minimize the effect of these outliers (Dixon & Yuen, 1974). The FTF internalizing and externalizing scales were not normally distributed, and therefore we performed a square root transformation to reduce skewness.

A preliminary correlation analysis was conducted to investigate the validity of constructs between FTF and SDQ. Reliable evidence of good construct validity was found, with strong correlations between FTF internalizing and SDQ emotion (r = .58, p < .01) and FTF externalizing and SDQ conduct (r = .57, p < .01) scales. Thus, when analyzing the study results, FTF internalizing and SDQ emotion scales are conceptualized as internalizing problems, and FTF externalizing and SDQ conduct scales as externalizing problems.

In order to investigate the nature of the relationship between parental postpartum bonding at eight months and child social-emotional problems at 5 years, separate linear regression analyses were conducted for mothers and fathers, and for each of the child outcome variables. First, the models included only maternal/paternal postpartum bonding as a predictor of child social-emotional problems. Secondly the models were adjusted for parental age, the number of previous children, parental education level, and child sex.

To investigate the joint effect of both parents’ postpartum bonding quality on child social-emotional problems, a maternal bonding x paternal bonding-interaction variable was analyzed in a model comprising also the main effects of mother’s and father’s bonding and adjusted for both parents’ background factors and child sex. In order to visualize interaction effects, maternal and paternal postpartum bonding variables were recoded into two-category variables with a cutoff point of ≥12 points.

Lastly, we conducted separate linear regression analyses for maternal and paternal bonding and child social-emotional problems and included parental depressive symptoms along with previously mentioned background factors.

## Results

### Descriptive statistics

Descriptive statistics for mothers, fathers and children are reported in Tables 1-2. The mean age for participating mothers was 31.2 years, and for fathers it was 32.8 years. 51% of mothers and 50% of fathers were first-time parents, and 17% of parents had two or more previous children. 36% of the mothers and 33% of the fathers had a university degree. Of the children in the study, 337 were girls (47.5%) and 373 were boys (52.5%).

**Table 1.** Descriptive statistics of maternal and paternal variables.

|  | Mothers |  | Fathers |  |
| --- | --- | --- | --- | --- |
| Categorical variables | N | % | N | % |
| Number of children (at baseline) |  |  |  |  |
| 0 | 333 | 51.1 | 302 | 50.4 |
| 1 | 211 | 32.4 | 193 | 32.2 |
| 2 or more | 108 | 16.6 | 104 | 17.4 |
| Education level (at baseline) |  |  |  |  |
| no degree / other | 46 | 6.5 | 72 | 10.7 |
| occupational / appl. univ. degree | 407 | 57.6 | 384 | 56.8 |
| university degree | 254 | 35.9 | 220 | 32.5 |
| Continuous variables | Mean | SD | Mean | SD |
| Age (at baseline) | 31.18 | 4.58 | 32.78 | 5.09 |
| Postpartum depression (CES-D) (at 8 months) | 5.42 | 3.99 | 4.23 | 3.64 |
| Postpartum bonding (PBQ) (at 8 months) | 4.24 | 3.65 | 5.06 | 4.12 |

**Table 2.** Descriptive statistics of child social-emotional outcome variables at age 5 years by child sex.

| Child outcome | Girls |  | Boys |  |
| --- | --- | --- | --- | --- |
|  | Mean | SD | Mean | SD |
| FTF internalizing problems | 1.51 | 0.50 | 1.49 | 0.50 |
| FTF externalizing problems | 1.75 | 0.71 | 1.84 | 0.79 |
| SDQ emotional problems | 1.49 | 1.49 | 1.28 | 1.45 |
| SDQ conduct problems | 1.87 | 1.50 | 2.05 | 1.65 |
| SDQ peer problems | 1.66 | 1.33 | 1.72 | 1.44 |

Regarding the associations between study and background variables, there were statistically significant moderate positive correlations between maternal postpartum depressive symptoms and maternal postpartum bonding difficulties (r = .51) and between paternal postpartum depressive symptoms and paternal postpartum bonding difficulties (r = .46).

Regarding the background factors, mothers’ higher number of children correlated with maternal postpartum bonding difficulties and higher child internalizing problems. Larger paternal number of children correlated with higher child internalizing and peer problems. There was an association between higher paternal education level and child’s lower level of peer problems. Lower maternal age correlated with higher child internalizing problems. Maternal and paternal postpartum depressive symptoms correlated with both child’s internalizing and externalizing problems, and paternal symptoms correlated also with child peer problems.

### Maternal postpartum bonding and child social-emotional problems

In the unadjusted linear regression model 1 (Table 3), maternal postpartum bonding difficulties were significantly associated with all the child social-emotional problem outcome variables. In the second model, adjustment of maternal and child background factors did not affect the results (Table 3).

**Table 3.** Regression results for maternal postpartum bonding as a predictor of child social-emotional problems (Model 1), controlled for maternal and child demographic factors (Model 2).

| Child outcome variable | Model 1 |  |  |  | Model 2 |  |  |  |
| --- | --- | --- | --- | --- | --- | --- | --- | --- |
| | $\beta$ | SE | p | R<sup>2</sup> | $\beta$ | SE | p | R<sup>2</sup> |
| FTF internalizing problems | .185 | .005 | <.001 | .033 | .174 | .005 | <.001 | .043 |
| FTF externalizing problems | .124 | .008 | .001 | .014 | .119 | .008 | .004 | .014 |
| SDQ emotional problems | .172 | .016 | <.001 | .028 | .175 | .016 | <.001 | .063 |
| SDQ conduct problems | .112 | .017 | .004 | .011 | .111 | .018 | .007 | .012 |
| SDQ peer problems | .119 | .015 | .002 | .013 | .120 | .016 | .004 | .010 |

Note. Bolded p-values are statistically significant, R<sup>2</sup>'s represent adjusted R<sup>2</sup> value.

### Paternal postpartum bonding and child social-emotional problems

In the unadjusted linear regression model 1 (Table 4), paternal postpartum bonding difficulties were associated with all the outcome variables except for SDQ emotional problems. In the background-adjusted model 2 (Table 4), paternal bonding was related to higher child internalizing and externalizing problems (FTF) as well as problems in peer relations (SDQ).

**Table 4.**
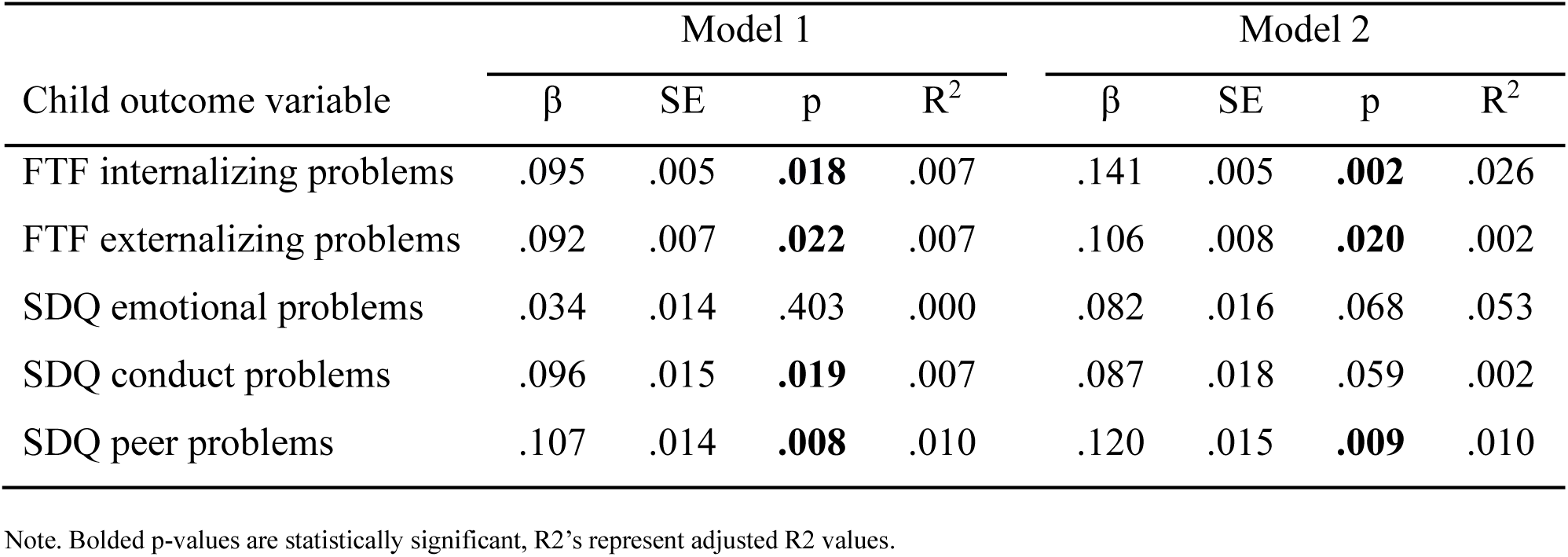
Regression results for paternal postpartum bonding as a predictor of child social-emotional problems (Model 1), controlled for paternal and child demographic factors (Model 2).

### The joint effect of maternal and paternal bonding on child social-emotional problems

Regarding the joint effect of maternal and paternal bonding quality on child social-emotional problems, there was one statistically significant Maternal bonding X Paternal bonding interaction term, indicating the association between the joint parental bonding quality and SDQ emotional problems (p = .004) (see Table A1 for the interaction terms in Appendix). This is visualized in Figure 1.

**Fig. 1.**
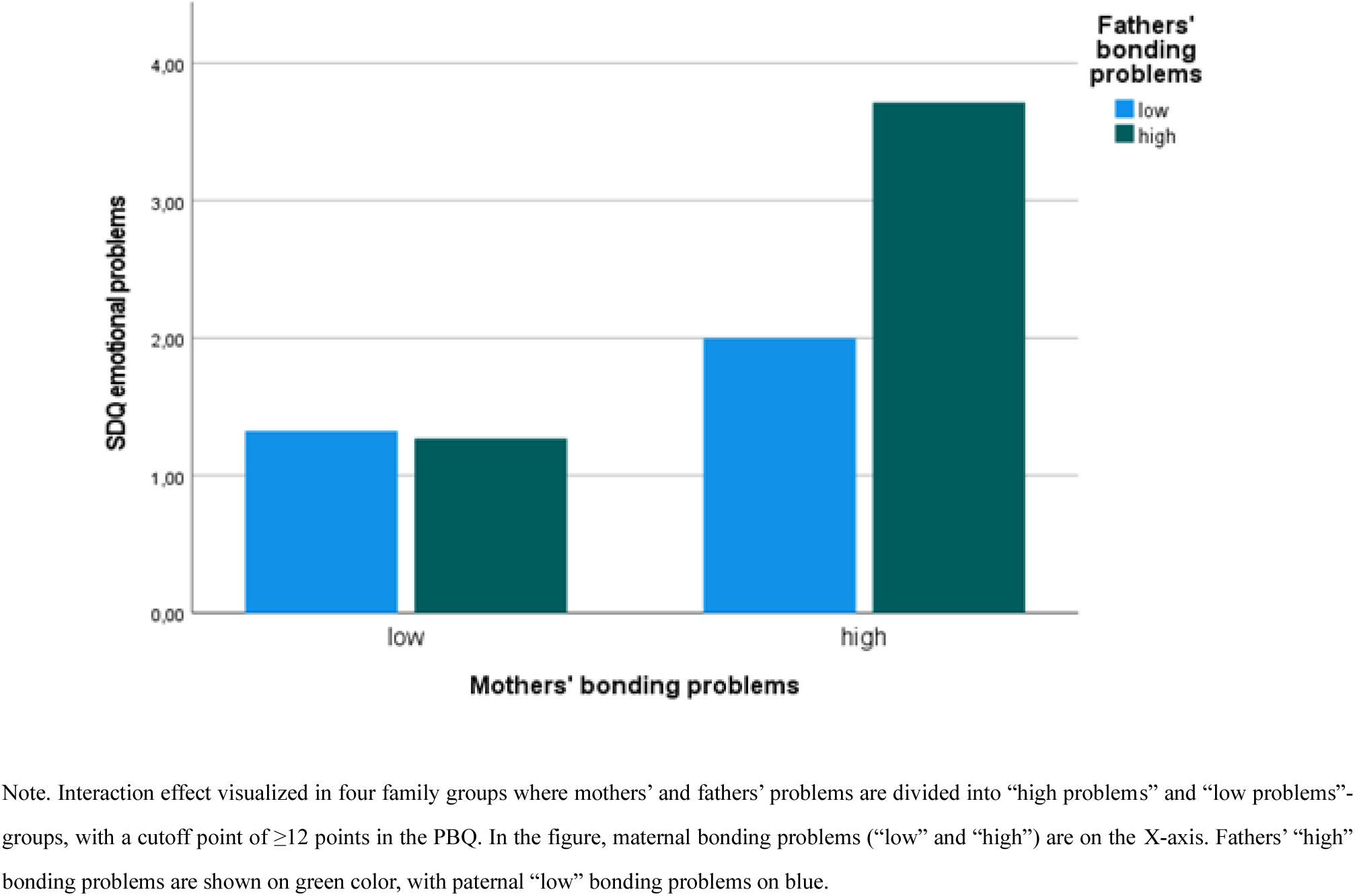
Child emotional problems by maternal and paternal bonding quality.

The figure shows that higher maternal bonding difficulties were associated with SDQ emotional problems irrespective of paternal bonding difficulties. Paternal bonding difficulties were associated with child emotional problems when mothers also had bonding difficulties. Higher amount of both paternal and maternal bonding difficulties posed the most significant risk for child emotional problems.

### Controlling depressive symptoms to study the association of parental bonding and child social-emotional problems

When adding maternal postpartum depressive symptoms to the model (Table 5), the relationship between postpartum maternal bonding and internalizing and peer problems remained significant. Yet, maternal bonding no longer predicted externalizing problems (FTF externalizing, SDQ conduct). Maternal postpartum depressive symptoms were associated with the FTF internalizing and externalizing problems (Table 5).

**Table 5.** Regression results for maternal postpartum bonding as a predictor of child social-emotional problems, controlled for demographic factors and maternal depressive symptoms.

| Child outcome variable | Coefficients for postpartum bonding |  |  | Coefficients for depressive symptoms |  |  | Model |
| --- | --- | --- | --- | --- | --- | --- | --- |
| | $\beta$ | SE | p | $\beta$ | SE | p | R<sup>2</sup> |
| FTF internalizing problems | .107 | .006 | **.027** | .124 | .006 | **.010** | .051 |
| FTF externalizing problems | .063 | .010 | .199 | .104 | .009 | **.033** | .020 |
| SDQ emotional problems | .155 | .014 | **.001** | .038 | .018 | .420 | .060 |
| SDQ conduct problems | .081 | .021 | .093 | .058 | .020 | .229 | .012 |
| SDQ peer problems | .113 | .019 | **.019** | .009 | .017 | .847 | .008 |

Note. Bolded p-values are statistically significant, R2's represent adjusted R2 values.

The relationship between paternal postpartum bonding and child internalizing problems only remained significant for FTF internalizing problems, when controlling for paternal depressive symptoms (Table 6). Paternal postpartum depressive symptoms were only associated with SDQ conduct problems (Table 6).

**Table 6.** Regression results for paternal postpartum bonding as a predictor of child social-emotional problems, controlled for demographic factors and paternal depressive symptoms.

| Child outcome variable | Coefficients for postpartum bonding |  |  | Coefficients for depressive symptoms |  |  | Model |
| --- | --- | --- | --- | --- | --- | --- | --- |
| | $\beta$ | SE | p | $\beta$ | SE | p | R<sup>2</sup> |
| FTF internalizing problems | .109 | .006 | **.032** | .069 | .007 | .171 | .028 |
| FTF externalizing problems | .068 | .009 | .186 | .089 | .010 | .081 | .007 |
| SDQ emotional problems | .077 | .018 | .130 | .011 | .020 | .831 | .051 |
| SDQ conduct problems | .031 | .020 | .552 | .127 | .022 | **.013** | .013 |
| SDQ peer problems | .089 | .018 | .085 | .069 | .019 | .175 | .012 |

Note. Bolded p-values are statistically significant, R2's represent adjusted R2 values.

## Discussion

The aim of this study was to investigate the individual and joint associations between maternal and paternal postpartum bonding on child social-emotional problems at preschool age. In line with our hypothesis, maternal postpartum bonding was associated with both internalizing, externalizing, and peer problems in child social-emotional development. Similar to mothers, paternal postpartum bonding was associated with later child social-emotional problems. Regarding joint effects, we found both cumulative and mother-driven effects: Maternal, but not paternal bonding problems alone predicted children’s internalizing problems, but families where both parents had bonding problems showed the worst outcome. Controlling for parental depressive symptoms diminished some of the effects.

As hypothesized, maternal postpartum bonding was associated with later child internalizing, externalizing and peer problems. The results are well in line with recent bonding studies in toddlers (e.g., Le Bas et al., 2022; Murakami et al., 2023; Rusanen et al., 2022), further advancing research by showing that the effects remain at least until five years of age. Our results thus suggest that dysfunctional maternal bonding is an important risk factor for later childhood social-emotional problems. In line with our hypothesis, also paternal postpartum bonding was associated with child social-emotional problems. This is a novel finding for postpartum bonding research, as there are no previous studies investigating fathers’ bonding and child social-emotional outcomes. Yet, there are studies on the impact of paternal mental wellbeing and involvement showing similar associations with child social-emotional development (e.g., Baker, 2017; Ramchandani et al., 2008). Hence, our results add to the existing literature by highlighting the importance of the paternal bond for later child development and wellbeing.

This was the first study investigating the joint effect of maternal and paternal postpartum bonding on later child social-emotional problems. In terms of child internalizing problems, the results showed that children whose mothers had bonding difficulties had more internalizing problems than children whose fathers had difficulties in bonding, when the other parent showed no bonding difficulties. The combination of both maternal and paternal bonding difficulties posed the highest risk for child internalizing problems, indicating both cumulative and mother-driven risk effects. Although fathers’ bonding difficulties alone did not seem to increase child internalizing problems, they aggravated the effect of maternal dysfunctional bonding on later child internalizing problems. Despite the lack of previous research, this finding can be supported by attachment and postpartum depression research that has often indicated similar cumulative and mother-driven effects. Studies have shown that paternal attachment quality and depressive symptoms alone may not greatly increase the risk of child internalizing problems, but the risk is accumulated when both parents have attachment difficulties or depressive symptoms (Cioffi et al., 2021; Kochanska & Kim, 2013; Pietikäinen et al., 2020). The current study thus expands the literature by showing that this also applies to parental bonding.

A common finding for mothers and fathers was that with both parents, externalizing problems seemed to be more strongly associated with parental depressive symptoms than with parental bonding quality. This is well in line with previous postpartum depression research findings showing that maternal and paternal postpartum depressive symptoms are a risk factor for later child externalizing problems (e.g., Kane & Garber, 2009; Kim-Cohen et al., 2005). Kane and Garber (2009) have suggested that the association between parental depression and child externalizing problems could be mediated by a higher tendency for parent-child conflict. In this study, only maternal postpartum bonding remained a significant predictor for later child internalizing and peer problems after controlling for depressive symptoms. The role of maternal bonding for later child internalizing and peer problem development could thus be somewhat distinct and broader than the role of paternal bonding. In this study, the majority of respondents for child questionnaires were mothers, which could alternatively explain the stronger association between maternal bonding and child internalizing and peer problems.

This study is from the large CHILD-SLEEP-cohort study sample representative of the Finnish general population (Paavonen et al., 2017), which can be considered a significant strength. One of the most central strengths of this study is its longitudinal nature with a long follow-up period. To our knowledge, there are no prior studies investigating the impact of postpartum bonding quality well beyond toddlerhood. Recently, Murakami and colleagues (2022) examined maternal postpartum bonding and child social-emotional problems at the age of four, and the current study extends one year further. Even more importantly, there is no previous knowledge of the impact of postpartum bonding on child social-emotional development from the perspective of both mothers and fathers. In attachment and bonding research, fathers have thus far been underrepresented, and more information is needed on the nature and importance of the developing emotional bond between the father and the child. Thirdly, parental postpartum bonding was examined alongside postpartum depression, which can be considered an advantage. Analyzing postpartum bonding and child social-emotional problems with and without the impact of parental depression enables a closer look to the partial overlap and differences of the two constructs, offering an important perspective to conceptualize the possible risk factors for early parent-child relationship.

There are also limitations to consider. In this study, self-reports were used, which generally increases probability for socially desirable and culturally accepted answering patterns (Zerbe & Paulhus, 1987). Parenting and mental health are both especially sensitive and personal subjects, and parents are prone to underestimate the gravity of issues in these areas (Bornstein et al., 2015). Thus, the levels of parental bonding difficulties and child social-emotional problems may in fact be higher than what the self-evaluation can capture. Another point to consider is the generalizability of results in the population level. With the long study period, there was a follow-up rate of 41% in the 5-year follow-up. The parents with higher education level, older maternal age and fewer children were over-represented in the current sample. Higher socioeconomic status has been associated with lower child social-emotional problems (e.g., Bradley & Corwyn, 2002), while lower maternal age is associated with higher childhood internalizing and externalizing problems (Edwards & Hans, 2015). Therefore, the results must be interpreted carefully, as they may be generalizable only to families with relatively good socioeconomic status. Another limitation concerns the relatively low reliability of measures, especially the Strengths and Difficulties Questionnaire (Goodman, 1997). The SDQ is a world widely used general child social-emotional questionnaire with satisfactory internal consistency in the British population sample (Goodman, 2001) but may not fit optimally with Finnish population sample used in our study (Kankaanpää et al., 2023). Yet, we used also the Five-to-Fifteen questionnaire by Kadesjö and colleagues (2004) which proved to have better internal consistency in our sample. The SDQ is shorter than the FTF questionnaire, which could partially explain weaker internal reliability of constructs.

In this study, several potentially significant parent-related factors were not considered. The length of parental leave may affect the parent-child relationship (Clark et al., 1997; Petts et al., 2020; Plotka & Busch-Rossnagel, 2018). According to Finnish statistics, fathers nowadays are using only 16 percent of family-allocated parental leave days (Keskinen, 2023), making paternity leaves significantly shorter than maternity leaves which can last up to around 300 parental leave days. Also, it is suggested that fathers’ bonding is affected more by romantic relationship satisfaction than mothers’ bonding (Morris et al., 2022). There is preliminary research suggesting that parental partner support is associated with toddler social-emotional development, mediated by postpartum bonding (de Waal et al., 2023). These family dynamic factors can explain some of the differences in the roles of maternal and paternal bonding for child social-emotional development and should be examined in future studies. Lastly, future research should also investigate the potential moderating or mediating associations of postpartum bonding and parental depressive symptoms in terms of child internalizing and externalizing behaviors.

### Summary

Early parent-child interactions are affected by several parent- and child-related factors, one of which is the quality of the emotional bond that parents form with the child. The concept of postpartum bonding is a growing research area with a potentially high practical importance for child development and mental health. The aim of this study was to investigate how parental postpartum bonding is associated with child social-emotional problems at the age of five and was the first to include fathers in the study. Based on this study, especially mothers’ early bonding difficulties may predict later child internalizing and social problems in preschool age, regardless of postpartum depression. However, the results also highlight the importance of both mothers’ and fathers’ bonding quality, as children are most susceptible to develop internalizing problems in families where both parents have dysfunctional bonding. Parental postpartum bonding should be one of the central subjects of discussion when targeting early risk factors for child wellbeing. In the future, it would be beneficial to screen for postpartum bonding difficulties for both mothers and fathers in maternity and childcare clinics to help detect potential risk groups for child mental health problems early on. Helping parents develop functional bonds with their children by means of psychoeducation or focused parenting interventions within the first year of life could greatly affect the developmental trajectories of children.

## Data availability

The data cannot be shared publicly due to legal restrictions (Finnish Data Protection Act 1050/2018) and the nature of the data (individual level data). Data are available upon request (contact). Data requests are reviewed in Finnish Institute for Health and Welfare for compliance with Finnish law, regulations, and ethical guidelines.

## Acknowledgements

This work was supported by the Academy of Finland (grant numbers 134880, 308588, 342747, 351492), the Gyllenberg Foundation, the Yrjö Jahnson Foundation, the Foundation for Pediatric Research, the Finnish Cultural Foundation.

## Author contribution

This study is part of the CHILD-SLEEP-cohort study by the Finnish Institute for Health and Welfare. The first version of the manuscript was drafted by Enni Hatakka. All authors participated in the critical revision of the manuscript and approved the final version of the manuscript.

## APPENDIX

**Table A1.** Regression results for the interaction effect of joint parental bonding quality and child social-emotional outcomes, controlled for maternal, paternal and child background factors.

| Child outcome | $\beta$ | SE | p | R<sup>2</sup> |
| --- | --- | --- | --- | --- |
| FTF internalizing problems | .088 | .001 | .373 | .072 |
| FTF externalizing problems | -.009 | .002 | .932 | .025 |
| SDQ emotional problems | .228 | .003 | **.018** | .088 |
| SDQ conduct problems | -.171 | .004 | .085 | .027 |
| SDQ peer problems | .167 | .003 | .090 | .037 |

## References

Baker, C. E. (2017). Father-son relationships in ethnically diverse families: Links to boys’ cognitive and social emotional development in preschool. Journal of Child and Family Studies, 26(8), 2335–2345.

Bieleninik, Ł., Lutkiewicz, K., Jurek, P., & Bidzan, M. (2021). Paternal postpartum bonding and its predictors in the early postpartum period: cross-sectional study in a polish cohort. Frontiers in Psychology, 12, 1112. 10.3389/fpsyg.2021.628650

Björgvinsson, T., Kertz, S. J., Bigda-Peyton, J. S., McCoy, K. L., & Aderka, I. M. (2013). Psychometric properties of the CES-D-10 in a psychiatric sample. Assessment, 20(4), 429–436.10.1177/1073191113481998

Bornstein, M. H., Putnick, D. L., Lansford, J. E., Pastorelli, C., Skinner, A. T., Sorbring, E., … & Oburu, P. (2015). Mother and father socially desirable responding in nine countries: Two kinds of agreement and relations to parenting self-reports. International Journal of Psychology, 50(3), 174–185. 10.1002/ijop.12084

Bradley, R. H., & Corwyn, R. F. (2002). Socioeconomic status and child development. Annual review of psychology, 53(1), 371–399. 10.1146/annurev.psych.53.100901.135233

Brandon, A. R., Pitts, S., Denton, W. H., Stringer, C. A., & Evans, H. (2009). A history of the theory of prenatal attachment. Journal of prenatal & perinatal psychology & health: APPPAH, 23(4), 201.

Brockington, I. F., Fraser, C., & Wilson, D. (2006). The postpartum bonding questionnaire: a validation. Archives of women’s mental health, 9(5), 233–242. 10.1007/s00737-006-0132-1

Cadman, T., Belsky, J., & Pasco Fearon, R. M. (2018). The Brief Attachment Scale (BAS-16): A short measure of infant attachment. Child: care, health and development, 44(5), 766–775. 10.1111/cch.12599

Campbell, S. B., Shaw, D. S., & Gilliom, M. (2000). Early externalizing behavior problems: Toddlers and preschoolers at risk for later maladjustment. Development and psychopathology, 12(3), 467–488. 10.1111/j.1469-7610.1995.tb01657.x

Cioffi, C. C., Leve, L. D., Natsuaki, M. N., Shaw, D. S., Reiss, D., Ganiban, J. M., & Neiderhiser, J. M. (2021). Examining reciprocal associations between parent depressive symptoms and child internalizing symptoms on subsequent psychiatric disorders: An adoption study. Depression and anxiety, 38(12), 1211–1224. 10.1002/da.23190

Clark, R., Hyde, J. S., Essex, M. J., & Klein, M. H. (1997). Length of maternity leave and quality of mother-infant interactions. Child development, 364–383. 10.2307/1131855

Condon, J. T. (1993). The assessment of antenatal emotional attachment: development of a questionnaire instrument. British journal of medical psychology, 66(2), 167–183. 10.1111/j.2044-8341.1993.tb01739.x

Condon, J. T., & Corkindale, C. J. (1998). The assessment of parent-to-infant attachment: Development of a self-report questionnaire instrument. Journal of Reproductive and Infant Psychology, 16(1), 57–76. 10.1080/02646839808404558

de Waal, N., Boekhorst, M. G., Nyklíček, I., & Pop, V. J. (2023). Maternal-infant bonding and partner support during pregnancy and postpartum: Associations with early child social-emotional development. Infant Behavior and Development, 72, 101871. 10.1016/j.infbeh.2023.101871

Dixon, W. J., & Yuen, K. K. (1974). Trimming and winsorization: A review. Statistische Hefte, 15(2-3), 157–170. 10.1007/bf02922904

Edwards, R. C., & Hans, S. L. (2015). Infant risk factors associated with internalizing, externalizing, and co-occurring behavior problems in young children. Developmental Psychology, 51(4), 489. https://psycnet.apa.org/doi/10.1037/a0038800

Figueiredo, B., & Costa, R. (2009). Mother’s stress, mood and emotional involvement with the infant: 3 months before and 3 months after childbirth. Archives of women’s mental health, 12, 143–153. 10.1007/s00737-009-0059-4

Fonseca, A., Nazaré, B., & Canavarro, M. C. (2018). Mothers’ and fathers’ attachment and caregiving representations during transition to parenthood: an actor–partner approach. Journal of Reproductive and Infant Psychology, 36(3), 246–260. 10.1080/02646838.2018.1449194

Goodman, R. (1997). The Strengths and Difficulties Questionnaire: a research note. Journal of child psychology and psychiatry, 38(5), 581–586. 10.1111/j.1469-7610.1997.tb01545.x

Goodman, R. (2001). Psychometric properties of the strengths and difficulties questionnaire. Journal of the American Academy of Child & Adolescent Psychiatry, 40(11), 1337–1345. 10.1097/00004583-200111000-00015

Hay, D. F., Payne, A., & Chadwick, A. (2004). Peer relations in childhood. Journal of child psychology and psychiatry, 45(1), 84–108. 10.1046/j.0021-9630.2003.00308.x

Kadesjö, B., Janols, L. O., Korkman, M., Mickelsson, K., Strand, G., Trillingsgaard, A., & Gillberg, C. (2004). The FTF (Five to Fifteen): the development of a parent questionnaire for the assessment of ADHD and comorbid conditions. European child & adolescent psychiatry, 13(3), iii3–iii13. 10.1007/s00787-004-3002-2

Kane, P., & Garber, J. (2009). Parental depression and child externalizing and internalizing symptoms: Unique effects of fathers’ symptoms and perceived conflict as a mediator. Journal of Child and Family Studies, 18, 465–472. 10.1007/s10826-008-9250-x

Kankaanpää, R., Töttö, P., Punamäki, R. L., & Peltonen, K. (2023). Is it time to revise the SDQ? The psychometric evaluation of the Strengths and Difficulties Questionnaire. Psychological Assessment. 10.1037/pas0001265

Keskinen, S. (2023). Kelan lapsiperhe-etuustilasto 2022. http://hdl.handle.net/10138/358139

Kim-Cohen, J., Moffitt, T. E., Taylor, A., Pawlby, S. J., & Caspi, A. (2005). Maternal depression and children’s antisocial behavior: nature and nurture effects. Archives of general psychiatry, 62(2), 173–181. 10.1001/archpsyc.62.2.173

Kochanska, G., & Kim, S. (2013). Early attachment organization with both parents and future behavior problems: From infancy to middle childhood. Child development, 84(1), 283–296.

Le Bas, G. A., Youssef, G. J., Macdonald, J. A., Rossen, L., Teague, S. J., Kothe, E. J., … & Hutchinson, D. M. (2020). The role of antenatal and postnatal maternal bonding in infant development: A systematic review and meta-analysis. Social development, 29(1), 3–20. 10.1111/sode.12392

Le Bas, G., Youssef, G., Macdonald, J. A., Teague, S., Mattick, R., Honan, I., … & Hutchinson, D. (2022). The role of antenatal and postnatal maternal bonding in infant development. Journal of the American Academy of Child & Adolescent Psychiatry, 61(6), 820–829. 10.1016/j.jaac.2021.08.024

Liu, J. (2004). Childhood externalizing behavior: Theory and implications. Journal of child and adolescent psychiatric nursing, 17(3), 93–103. 10.1111/j.1744-6171.2004.tb00003.x

McNamara, J., Townsend, M. L., & Herbert, J. S. (2019). A systemic review of maternal wellbeing and its relationship with maternal fetal attachment and early postpartum bonding. PloS one, 14(7), e0220032. 10.1371/journal.pone.0220032

Mesman, J., Bongers, I. L., & Koot, H. M. (2001). Preschool developmental pathways to preadolescent internalizing and externalizing problems. The Journal of Child Psychology and Psychiatry and Allied Disciplines, 42(5), 679–689.

Mohebbi, M., Nguyen, V., McNeil, J. J., Woods, R. L., Nelson, M. R., Shah, R. C., … & ASPREE Investigator Group. (2018). Psychometric properties of a short form of the Center for Epidemiologic Studies Depression (CES-D-10) scale for screening depressive symptoms in healthy community dwelling older adults. General hospital psychiatry, 51, 118–125. 10.1016/j.genhosppsych.2017.08.002

Moore, S. E., Norman, R. E., Suetani, S., Thomas, H. J., Sly, P. D., & Scott, J. G. (2017). Consequences of bullying victimization in childhood and adolescence: A systematic review and meta-analysis. World journal of psychiatry, 7(1), 60.

Morris, A. R., Aviv, E. C., Khaled, M., Corner, G. W., & Saxbe, D. E. (2022). Prenatal romantic relationship satisfaction predicts parent–infant bonding for fathers, but not mothers. Couple and Family Psychology: Research and Practice. https://psycnet.apa.org/doi/10.1037/cfp0000225

Murakami, K., Ishikuro, M., Obara, T., Noda, A., Ueno, F., Onuma, T., … & Kuriyama, S. (2023). Maternal postnatal bonding disorder and emotional/behavioral problems in preschool children: The Tohoku Medical Megabank Project Birth and Three-Generation Cohort Study. Journal of Affective Disorders. 10.1016/j.jad.2023.01.044

Paavonen, E. J., Saarenpää-Heikkilä, O., Pölkki, P., Kylliäinen, A., Porkka-Heiskanen, T., & Paunio, T. (2017). Maternal and paternal sleep during pregnancy in the Child-sleep birth cohort. Sleep medicine, 29, 47–56. 10.1016/j.sleep.2016.09.01

Petts, R. J., Knoester, C., & Waldfogel, J. (2020). Fathers’ paternity leave-taking and children’s perceptions of father-child relationships in the United States. Sex Roles, 82, 173–188. 10.1007/s11199-019-01050-y

Pietikäinen, J. T., Kiviruusu, O., Kylliäinen, A., Pölkki, P., Saarenpää-Heikkilä, O., Paunio, T., & Paavonen, E. J. (2020). Maternal and paternal depressive symptoms and children’s emotional problems at the age of 2 and 5 years: a longitudinal study. Journal of Child Psychology and Psychiatry, 61(2), 195–204. 10.1111/jcpp.13126

Plotka, R., & Busch-Rossnagel, N. A. (2018). The role of length of maternity leave in supporting mother–child interactions and attachment security among American mothers and their infants. International Journal of Child Care and Education Policy, 12(1), 1–18. 10.1186/s40723-018-0041-6

Radlof, L. S. (1977). The CES-D scale: A self report depression scale for research in the general population. Applied Psychological Measurement, 1(3), 385–401. 10.13072/midss.120

Ramchandani, P. G., Stein, A., O’CONNOR, T. G., Heron, J. O. N., Murray, L., & Evans, J. (2008). Depression in men in the postnatal period and later child psychopathology: a population cohort study. Journal of the American Academy of Child & Adolescent Psychiatry, 47(4), 390–398.

Rusanen, E., Lahikainen, A. R., Vierikko, E., Pölkki, P., & Paavonen, E. J. (2022). A Longitudinal Study of Maternal Postnatal Bonding and Psychosocial Factors that Contribute to Social-Emotional Development. Child Psychiatry & Human Development, 1-13. 10.1007/s10578-022-01398-5

Rusanen, E., Vierikko, E., Kojo, T., Lahikainen, A. R., Pölkki, P., & Paavonen, E. J. (2021). Prenatal expectations and other psycho-social factors as risk factors of postnatal bonding disturbance. Infant mental health journal, 42(5), 655–671. 10.1002/imhj.21941

Stacks, A. M. (2005). Using an ecological framework for understanding and treating externalizing behavior in early childhood. Early Childhood Education Journal, 32(4), 269–278. 10.1007/s10643-004-0754-8

Van Bussel, J. C., Spitz, B., & Demyttenaere, K. (2010). Three self-report questionnaires of the early mother-to-infant bond: reliability and validity of the Dutch version of the MPAS, PBQ and MIBS. Archives of women’s mental health, 13(5), 373–384. 10.1007/s00737-009-0140-z

Verschueren, K., & Marcoen, A. (1999). Representation of self and socioemotional competence in kindergartners: Differential and combined effects of attachment to mother and to father. Child development, 70(1), 183–201. 10.1111/1467-8624.00014

Vreeswijk, C. M., Maas, A. J., Rijk, C. H., & van Bakel, H. J. (2014). Fathers’ experiences during pregnancy: Paternal prenatal attachment and representations of the fetus. Psychology of Men & Masculinity, 15(2), 129. https://psycnet.apa.org/doi/10.1037/a0033070

Vreeswijk, C. M., Rijk, C. H., Maas, A. J. B., & Van Bakel, H. J. (2015). Fathers’ and mothers’ representations of the infant: associations with prenatal risk factors. Infant Mental Health Journal, 36(6), 599–612. 10.1002/imhj.21541

Zerbe, W. J., & Paulhus, D. L. (1987). Socially desirable responding in organizational behavior: A reconception. Academy of management review, 12(2), 250–264. 10.5465/amr.1987.4307820

